# Plasma proteome signatures in sickle cell anaemia and effect of hydroxyurea treatment

**DOI:** 10.64898/2026.02.08.26345875

**Authors:** Neha Kumari, Sumit Paliwal, Anushri Umesh, Gopinath P, Jayalakshmi Marneni, Sristi Chakroborty, B Raman, Y Kameswari, K Ranjith Kumar, Suraj Nongmaithem, PSKDB Punyasri, Pradeep K Patra, Dipty Jain, Swasti Raychaudhuri, Giriraj Ratan Chandak

**Author notes:** Joint first authors (Neha Kumari, Sumit Paliwal).

## Abstract

**Background:** Sickle Cell Anaemia (SCA), a genetic blood disorder caused by a single mutation in the beta globin gene, displays a highly variable clinical course. Hydroxyurea (HU), an effective treatment, has an unclear mechanism of action. Plasma proteins can act as biomarkers for understanding disease states and response to HU treatment in SCA patients.

**Methods:** Plasma proteome profiling of 31 healthy individuals and 76 SCA patients, including those with and without HU treatment, was performed using a high-performance liquid chromatography system and Orbitrap mass spectrometer. Statistical analysis was performed to identify differentially abundant proteins (DAPs) between SCA patients and healthy controls. Subgroup analyses were performed to look at the impact of HU treatment on plasma proteome.

**Results:** Our analysis yielded 43 DAPs in the plasma of SCA patients. Global correlation and protein-protein network analysis revealed that these proteins are part of a robust interaction network. Proteins showing higher abundance (LBP, ORM1 and TFRC) were primarily associated with immune response whereas those with reduced abundance (FBLN1 and F13B) were linked to blood coagulation and proteolysis. Differential abundance of several proteins such as CD14, FCN3, LFALS3BP, LAP and TGFBI was observed in either male or female patients indicating influence of gender. Importantly, HU treatment was associated with elevated levels of haptoglobin (HP) and hemopexin (HPX), key proteins involved in free hemoglobin scavenging. Notably, DAPs such as F10, LPA, and FCN3 overlapped with proteins previously reported to be differentially abundant in beta-thalassemia patients. Moreover, multiple proteins, including APOL1, AZGP1, FBLN1, GPLD1, HPX, LGALS3BP, and TFRC correlated with clinical parameters, such as blood transfusion frequency and, vaso-occlusive crisis, and WBC and platelet counts.

**Conclusions:** This study identifies novel differentially abundant plasma proteins in SCA, expanding the current repertoire of disease-associated biomarkers and proteins modulated by hydroxyurea therapy. The observed overlap with beta-thalassemia associated signatures reinforces shared pathophysiological mechanisms between these hemoglobinopathies. Several of these proteins show significant correlations with key clinical parameters and disease complications, offering insights into disease mechanisms and potential utility in disease management. Collectively, these findings provide a strong foundation for translational validation in larger, independent cohorts.

## INTRODUCTION

Sickle Cell Anaemia (SCA) is a monogenic blood disorder caused by transversion of A to T (glutamic acid to valine) in sixth codon of the beta globin (HBB) gene that confers a sickle shape to red blood cells under hypoxic conditions (Chakravorty and Williams, 2015; GBD 2021 Sickle Cell Disease Collaborators, 2023). SCA is more prevalent in developing and under-developed countries of low-socio economic status with India having the second-highest burden after Sub-Saharan Africa (Piel *et al*., 2010; Brousse and Rees, 2021; GBD 2021 Sickle Cell Disease Collaborators, 2023). Over the past decades, Hydroxyurea (HU) has emerged as a potent, disease-modifying drug for SCA with best outcomes reported in younger patients (Power-Hays and Ware, 2020). However, the clinical course of SCA is highly variable and the precise molecular mechanisms underlying the disease and therapeutic efficacy of HU are still unclear. It is proposed that HU exerts its therapeutic effects by reversibly inhibiting ribonucleotide reductase, affecting DNA synthesis and repair, temporarily arresting hematopoiesis, altering erythroid kinetics, and thus increasing fetal haemoglobin (HbF) through recruitment of early erythroid progenitors (Elford, 1968; Bridges *et al*., 1996; Baliga *et al*., 2000; Koç *et al*., 2004; Alvino *et al*., 2007). Furthermore, HU therapy may also deregulate expression of surface molecules that adhere to the endothelium, ameliorate chronic inflammation, and elevate nitric oxide and cyclic nucleotides to facilitate vascular dilatation (Bridges *et al*., 1996; Cokic *et al*., 2003; McGann and Ware, 2015).

Circulatory proteins in blood plasma often surrogate for an individual’s health-state. Relative abundance of plasma proteins is regarded as markers for disease susceptibility, activity, or response to specific drugs and therefore widely used for prognostic and diagnostic purposes (Hu, Loo and Wong, 2006). Given the complex pathophysiology of SCA, identifying biomarkers for various disease states, response to HU-therapy etc. is vital for better disease management. Mass spectrometry based high-throughput, quantitative proteomics of blood plasma and serum can help in identifying potential biomarkers, but the workflows are intrinsically challenging due to highly dynamic range of protein abundances (Hortin and Sviridov, 2010). The challenges are multifaceted for SCA plasma proteomics in India, primarily stemming from difficulties in blood sample collection in remote areas, proper storage and subsequent transportation to the laboratories and separation of plasma from the transported blood. Consequently, reports on mass spectrometry based extensive plasma proteomics of SCA is scarce in literature and elaborative plasma proteomics effort using Indian SCA patient cohort has not been reported till-date.

Here, we summarize the results of a comparative plasma proteomics effort for Indian SCA patients across various age groups, gender, and treatment states. Our findings, in addition to confirming the status of previously identified plasma proteins as SCA biomarkers, identifies 22 novel potential biomarkers influenced by HU therapy, gender, or age.

## METHODS

### Study subjects

Patients and healthy individuals included in this study were recruited through the outpatient department in a hospital-based setting and through general population screening under the CSIR-Sickle Cell Anaemia Mission. In the mission, initial screening for SCA was done using solubility test followed by confirmation through Hemoglobin electrophoresis or HPLC. Finally, for all the individuals, the SCA status was confirmed by targeted genetic testing. Written informed consent was obtained from all the individuals for blood sample collection. For individuals under 18 years of age, the consent was provided by parents or the legal guardian.

### Plasma sample preparation and protein quantitation

Two millilitres of venous blood was collected in an EDTA-vacutainer and allowed to stand at room temperature (RT) for 30 min. Subsequently, the blood sample was transferred to a 2 ml microcentrifuge tube and centrifuged at 4,000 RPM for 20 minutes at 4°C. The clear plasma was transferred into a fresh 1.5 ml screw-cap microcentrifuge tube and stored at -80°C/-20°C for future use.

The total plasma protein concentration was measured using Amido-black assay (Dieckmann-Schuppert and Schnittler, 1997). For the assay, the plasma samples were first diluted to a ratio of 1:10 in Phosphate Buffered Saline. Next, 2μl and 4μl of diluted plasma from each sample was spotted in duplicate on a nitrocellulose membrane (Amersham Biosciences). As a standard, Bovine Serum Albumin (BSA) (G Biosciences) was spotted in linear concentration of 1μg/μl to 10μg/μl and the membrane was air-dried. The protein spots on the membrane were stained using Amido-Black staining solution (0.1% Amido-Black 10B (Merck), 45% methanol, 10% glacial acetic acid) for 10 minutes. Then, the membrane was destained 3-4 times using a destaining solution (90% methanol, 2% glacial acetic acid) to wash out the background dye and allowed to air-dry at RT. The dried stained spots were excised, placed into a micro-centrifuge tube containing elution buffer (50% ethanol, 50 mM NaOH, 0.1 mM EDTA) and incubated at 25°C for an hour on Thermomixer (Eppendorf) at 1200 RPM. Lastly, the absorbance of eluted protein bound dye was recorded at 630 nm using a multiplate reader (PerkinElmer EnSpire) (https://doi.org/10.1007/s42485-022-00096-z) and the BSA standard plot was used for estimating the plasma protein concentration.

### Sample preparation for rapid plasma proteomics

For each sample, 200 μg protein was reduced in (1:10 v/v) 8M urea, 7.5 mM Dithiothreitol (DTT) for 10 min at RT. Next, proteins were further reduced with 45mM DTT in 50 mM ammonium bi-carbonate buffer (1:5 v/v) by incubating at RT for 20 min. Then, the reduced proteins were alkylated by adding 100 mM Iodoacetamide (IAA) in 50 mM ammonium bicarbonate buffer in the same volume as 45mM DTT for 30 min at RT in the dark. Subsequently, the sample was diluted 10-fold with 50 mM ammonium bicarbonate buffer and in-solution digestion was performed with MS grade Trypsin (1:20, 15 ng/μl in 25mM ammonium bi-carbonate/ 1mM calcium chloride) for 16 hours at 37°C. Proteolysis was stopped by adding Trifluoroacetic acid (TFA, 10%), and digested plasma was vacuum dried in a concentrator (Eppendorf). The peptide mixture obtained was re-suspended in 5% Acetonitrile (ACN)/ 0.1% TFA. Lastly, 8 μg of peptide was desalted on 10 μl Pierce C18 column tips (Thermo Scientific) and vacuum dried.

### Liquid chromatography tandem mass spectrometry (LC-MS/MS)

The LC-MS/MS analysis of technical duplicate of each sample was performed on an EASY-nLC 1200 high-performance liquid chromatography system (Thermo Fisher Scientific) interfaced to an Orbitrap Exploris 240 hybrid quadrupole-orbitrap mass spectrometer (Thermo Fisher Scientific). The samples were loaded onto in-line Acclaim PepMap 100 (100 µm x 2 cm length; 5 µm particle size; 100Å pore size) (Thermo Fisher Scientific) trap column and separated using an analytical reverse-phase column of PepMapTM RSLC C18 (75 μm × 25 cm length; 2 μm particle size; 100Å pore size) (Thermo Fisher Scientific). . The total run time was for 120 min. Separation of plasma peptides was achieved using a linear gradient of 5% ACN, 0.2% formic acid (Solvent A) and 90% ACN, 0.2% formic acid (Solvent B) at a flow rate of 200 nL/min. The mass spectrometer was operated in a data–dependent acquisition (DDA) mode with Full scan mass range of 400-1650 *m/z* with resolution set to 60,000 at 200 *m/z*. The HCD-MS^2^ spectra were obtained using the application method of peptide with maximum injection time set to “auto”. The dynamic exclusion mode was set to “auto” with an acquisition cycle time fixed at 2 sec.

### Data processing

Proteome Discoverer (2.5.0.400 version, Thermo Fisher Scientific,) served as the interface for data processing of mass spectrometer generated raw files. Protein identification and quantification was performed against the human UniProtKB/Swiss-Prot database (version July 2023; release-2024-03/) with known contaminants (SequestHT search engine). The proteolytic enzyme selected was trypsin with a maximum allowance of two missed cleavages and a minimum peptide length of six amino acids. Variable modifications of methionine by oxidation and carbamidomethylation of cysteine were designated as fixed modifications. Precursor mass tolerances for peptide and fragment ion were set at 20 ppm and 0.5 Da, respectively. Other parameters included false discovery rate (FDR) of 1% for both protein and peptide, minimum 1 peptide to report identification. Label Free Quantification (LFQ) was performed using the Minora Feature Detector node in the processing workflow, the Precursor Ions Quantifier node and the Feature Mapper in the consensus workflow.

### Data Analysis

Following the data processing, only proteins with a minimum of two peptides and quantification in both technical replicates were retained. Further, proteins not quantified in more than 30% of samples were excluded from downstream statistical analyses. The data were log2 transformed, and median-centered normalization was conducted at the protein level across the samples using proBatch package (Čuklina *et al*., 2021). Missing values were imputed using the missForest algorithm (Stekhoven and Bühlmann, 2012; Jin *et al*., 2021) in R (version 4.3.2). Differentially abundant proteins (DAPs) between disease and control groups were identified using a non-parametric unpaired two-sided Welch t-test, with a fold change (FC) threshold of |FC| > 0.75 and a p-value ≤ 0.05, as determined by Perseus software (version 1.6.10.43). For the sake of convenience, terms higher/increased abundance and upregulation have been used interchangeably throughout the text. Similarly, terms lower/reduced abundance and downregulation have been used interchangeably. DAPs were visualized using volcano plots with the EnhancedVolcano package (https://github.com/kevinblighe/EnhancedVolcano) and principal component analysis (PCA) plots were created using the PCAtools package (https://github.com/kevinblighe/PCAtools), generated in R (version 4.3.2).

### Data availability

The data has been uploaded in the PRIDE (Vizcaíno *et al*., 2016) (https://www.ebi.ac.uk/pride) partner repository of the ProteomeXchange Consortium (https://www.proteomexchange.org), with the dataset identifier PXD062365.

### Protein set enrichment analysis

Gene Ontology (GO) and enrichment analyses were conducted using ShinyGO (Ge, Jung and Yao, 2020) (v.0.82) to explore the functional characteristics and biological relevance of the identified proteins. The analysis focused on the Biological Process (BP) domain to identify enriched pathways and processes, aiding the understanding of the underlying molecular mechanisms.

### Gender and age based analysis

An Analysis of Variance (ANOVA) was conducted to compare the relative abundance of DAPs in males and females across healthy controls and SCA patients. Likewise, to assess the impact of age, SCA patients were categorised into three age-groups: Children (0-12 years), Adolescents (13-18 years) and Adults (above 18 years) ANOVA was performed for multigroup comparison. Tukey’s post-hoc test was employed to evaluate pairwise comparisons and an adjusted p-value ≤ 0.05 was deemed statistically significant.

## RESULTS

### Study subjects

The details and distribution of the study subjects are provided in Table 1. The genetic status of all the study subjects for the sickle cell anaemia mutation was confirmed by Sanger Sequencing.

**Table 1:** Distribution of sickle cell anaemia patients and healthy individuals included in the study.

|  |  | **SCA Patients** |  |  |
| --- | --- | --- | --- | --- |
|  | **Healthy Controls** | **w/o-HU** | **w/-HU** | **Total** |
| **Number of Samples** | 31 | 41 | 35 | 76 |
| **Age range (years) (Median)** | 11-25 (14) | 0.3-39 (8.7) | 0.5-40 (10.4) | 0.3-40 (9.35) |
| Age Group 1 (0-12 years) | 10 | 27 | 22 | 49 |
| Age Group 2 (13-18 years) | 16 | 8 | 7 | 15 |
| Age Group 3 (>18 years) | 5 | 6 | 6 | 12 |
| **Gender** |  |  |  |  |
| Male | 14 (45.17%) | 21 (51.22%) | 21 (60%) | 42 (55.26%) |
| Female | 17 (54.83%) | 20 (48.78%) | 14 (40%) | 34 (44.74%) |

SCA, sickle cell anaemia; w/o-HU, without Hydroxyurea; w/-, with Hydroxyurea

At the time of enrollment, the median age of healthy controls was notably higher compared to the SCA patients. Out of the 76 SCA patients, 35 were undergoing hydroxyurea (HU) treatment (w/-HU) at the time of plasma sample collection (**Table 1**), because of a history of significantly higher episodes of vaso-occlusive crisis (VOC) [5.55 ± 5.37 vs 2.22 ± 1.81; p =0.006] and higher frequency of blood transfusion [8.32 ± 6.87 vs 3.50 ± 3.76; p = 0.004] compared to other SCA patients (**Table 2, Supplementary Figure 1**). HU-treated patients exhibited significantly higher Mean Corpuscular Volume (MCV) [90.89 ± 11.91 vs 84.72 ± 10.81; p = 0.047] and Mean Corpuscular Hemoglobin Concentration (MCHC) [33.24 ± 1.86 vs 32.00 ± 1.79; p = 0.014] but lower White Blood Cell (WBC) and platelet count. Further, HU-treated patients had significantly higher serum albumin levels (4.05 ± 0.51 vs 3.62 ± 0.38; p = 0.004) and lower alanine aminotransferase (ALT) levels [27.55 ± 19.42 vs 38.48 ± 17.94; p = 0.032] (**Table 2**).

**Table 2:** Clinical characteristics of sickle cell anaemia patients with- and without- Hydroxyurea treatment.

|  | w/o-HU |  |  | w/-HU |  |  |
| --- | --- | --- | --- | --- | --- | --- |
| Variable | N | Mean ± sd | | N | Mean ± sd | p-value |
| Vaso-occlusive crisis | 23 | 2.22 ± 1.81 | | 33 | 5.55 ± 5.37 | 0.006 |
| Blood Transfusion | 22 | 3.50 ± 3.76 | | 28 | 8.32 ± 6.87 | 0.004 |
| Acute Febrile Illness (AFI)-Infection | 23 | 2.74 ± 2.53 | | 26 | 4.00 ± 3.03 | Ns |
| **Complete Blood Count** |  |  |  |  |  |  |
| Red Blood Cells (RBC) ($\times 10^6$) | 18 | 3.32 ± 1.63 | | 31 | 3.13 ± 0.74 | ns |
| Hemoglobin (HB) (g/dL) | 24 | 8.05 ± 1.59 | | 35 | 8.42 ± 2.58 | ns |
| Mean Corpuscular Volume (MCV) ($\mu\text{m}^3$) | 25 | 84.72 ± 10.81 | | 33 | 90.89 ± 11.91 | 0.047 |
| Mean Corpuscular Hemoglobin (MCH) (pg) | 25 | 27.15 ± 4.14 | | 33 | 30.01 ± 4.32 | 0.014 |
| Mean Corpuscular Hemoglobin Concentration (MCHC) (g/dL) | 25 | 32.00 ± 1.79 | | 33 | 33.24 ± 1.86 | 0.014 |
| Red Cell Distribution Width (RDW) ($\mu\text{m}^3$) | 23 | 17.74 ± 3.07 | | 29 | 16.97 ± 1.90 | ns |
| Reticulocyte (%) | 25 | 5.54 ± 3.96 | | 28 | 4.30 ± 2.02 | ns |
| White Blood Cell (WBC) ($\times 10^3/\mu\text{L}$) | 25 | 13.44 ± 5.92 | | 35 | 8.93 ± 4.31 | 0.001 |
| Neutrophils (%) | 18 | 52.51 ± 11.99 | | 27 | 49.15 ± 9.69 | ns |
| Lymphocytes (%) | 20 | 40.98 ± 10.79 | | 27 | 43.93 ± 9.45 | ns |
| Eosinophils (%) | 17 | 2.22 ± 0.94 | | 26 | 2.35 ± 0.85 | ns |
| Monocytes (%) | 18 | 14.72 ± 46.75 | | 27 | 4.30 ± 5.80 | ns |
| Platelet ($\times 10^3/\mu\text{L}$) | 25 | 466.40 ± 242.46 | | 35 | 302.11 ± 161.13 | 0.003 |
| **Hemoglobin** |  |  |  |  |  |  |
| HbA | 25 | 1.67 ± 0.28 | | 29 | 3.19 ± 4.31 | ns |
| HbS | 25 | 76.82 ± 6.03 | | 32 | 74.16 ± 6.90 | ns |
| HbF | 25 | 18.16 ± 6.64 | | 32 | 18.99 ± 5.45 | ns |
| **Kidney-Function test** |  |  |  |  |  |  |
| Blood-Urea | 25 | 17.37 ± 6.98 | | 34 | 14.68 ± 9.10 | ns |
| Serum-Creatinine | 25 | 0.49 ± 0.09 | | 34 | 0.54 ± 0.13 | ns |
| Serum-Sodium | 23 | 136.83 ± 2.50 | | 19 | 136.19 ± 3.04 | ns |
| Serum-Potassium | 24 | 4.32 ± 0.68 | | 19 | 4.47 ± 0.59 | ns |
| Urine-Microalbumin | 22 | 10.99 ± 4.23 | | 21 | 8.20 ± 5.23 | ns |
| **Liver-Function test** |  |  |  |  |  |  |
| Serum-Bilirubin | 24 | 3.92 ± 2.96 | | 33 | 2.95 ± 1.94 | ns |
| Serum-albumin | 19 | 3.62 ± 0.38 | | 24 | 4.05 ± 0.51 | 0.004 |
| Aspartate Aminotransferase (AST) | 25 | 44.24 ± 12.97 | | 33 | 43.70 ± 21.66 | ns |
| Alanine Aminotransferase (ALT) | 25 | 38.48 ± 17.94 | | 33 | 27.55 ± 19.42 | 0.032 |
| Alkaline phosphatase (ALP) | 25 | 115.88 ± 28.60 | | 33 | 141.14 ± 64.56 | ns |
| Lactate Dehydrogenase (LDH) | 24 | 861.00 ± 191.61 | | 18 | 747.17 ± 221.45 | ns |

### Proteomic analysis

#### Comparative plasma proteome profiling of SCA patients and healthy controls

The study design and analysis plan is depicted in Figure 1. Rapid plasma proteome profiling was performed on samples from 31 healthy controls and 76 SCA patients including 16 patients for whom plasma proteome was analyzed both before and after HU treatment. However, for these 16 patients, only plasma proteome data before HU treatment was used for comparative proteome analysis.

**Figure 1:**
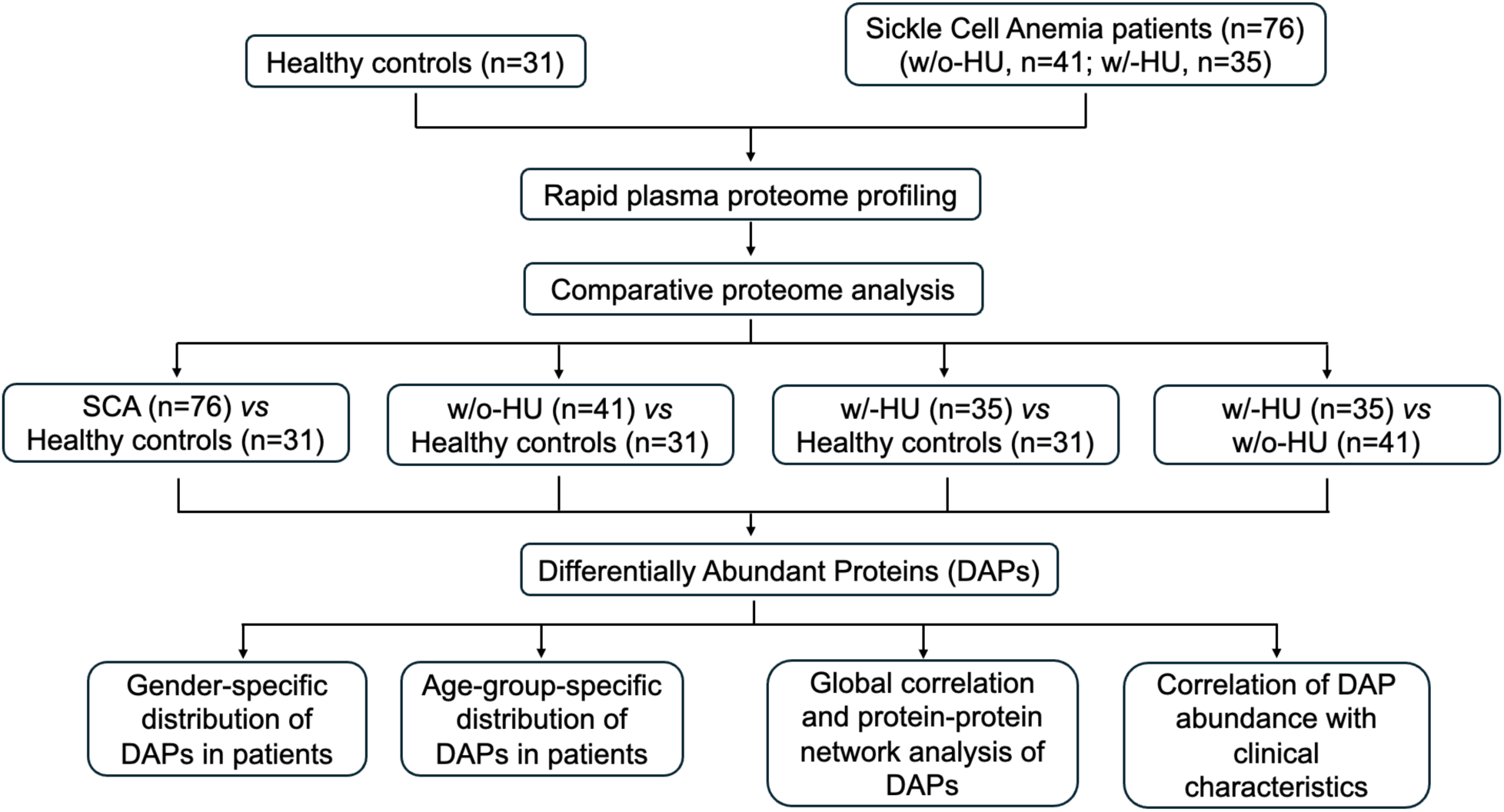
Schematic representation of study design and analysis plan for comparative plasma proteome profiling of sickle cell anaemia (SCA) patients and healthy controls. HU, Hydroxyurea; w/o-HU, without HU treatment; w/-HU, on HU treatment.

We quantified a total of 5969 peptides representing 432 proteins, of which 306 proteins were common between the healthy and SCA groups (**Fig. 2A**). On average, 200 proteins were quantified from individual plasma samples (**Fig. 2B**). The number of proteins quantified in healthy individuals ranged from 183 to 236 while the range was significantly higher, from 186 to 298, for SCA patients, indicating that the disease manifested a heterogeneous plasma proteome (**Fig. 2C**). When considering the criteria of presence of a protein in at least 70% of the samples, only 177 proteins were found to be common in both the groups. PCA with these 177 commonly quantified proteins revealed distinct plasma proteome composition between the groups (**Fig. 2D**). Comparative proteome analysis revealed 28 DAPs wherein 10 proteins showed reduced abundance in the plasma of SCA patients, while 18 proteins showed increased abundance (**Fig. 2E, Supplementary Table S1**). GO analysis revealed that proteins like Fibulin-1 (FBLN1) and coagulation factor XIII B subunit (F13B) which exhibited reduced abundance in SCA patients were linked to coagulation, fibrin clot formation, and proteolysis whereas proteins such as Lipopolysaccharide-binding protein (LBP), orosomucoid 1 (ORM1) and Transferrin Receptor 1 (TFRC) which displayed higher abundance were primarily involved in immune response regulation and influenced TNF signaling pathway (**Fig. 2F, Supplementary Figure 2**).

**Figure 2:**
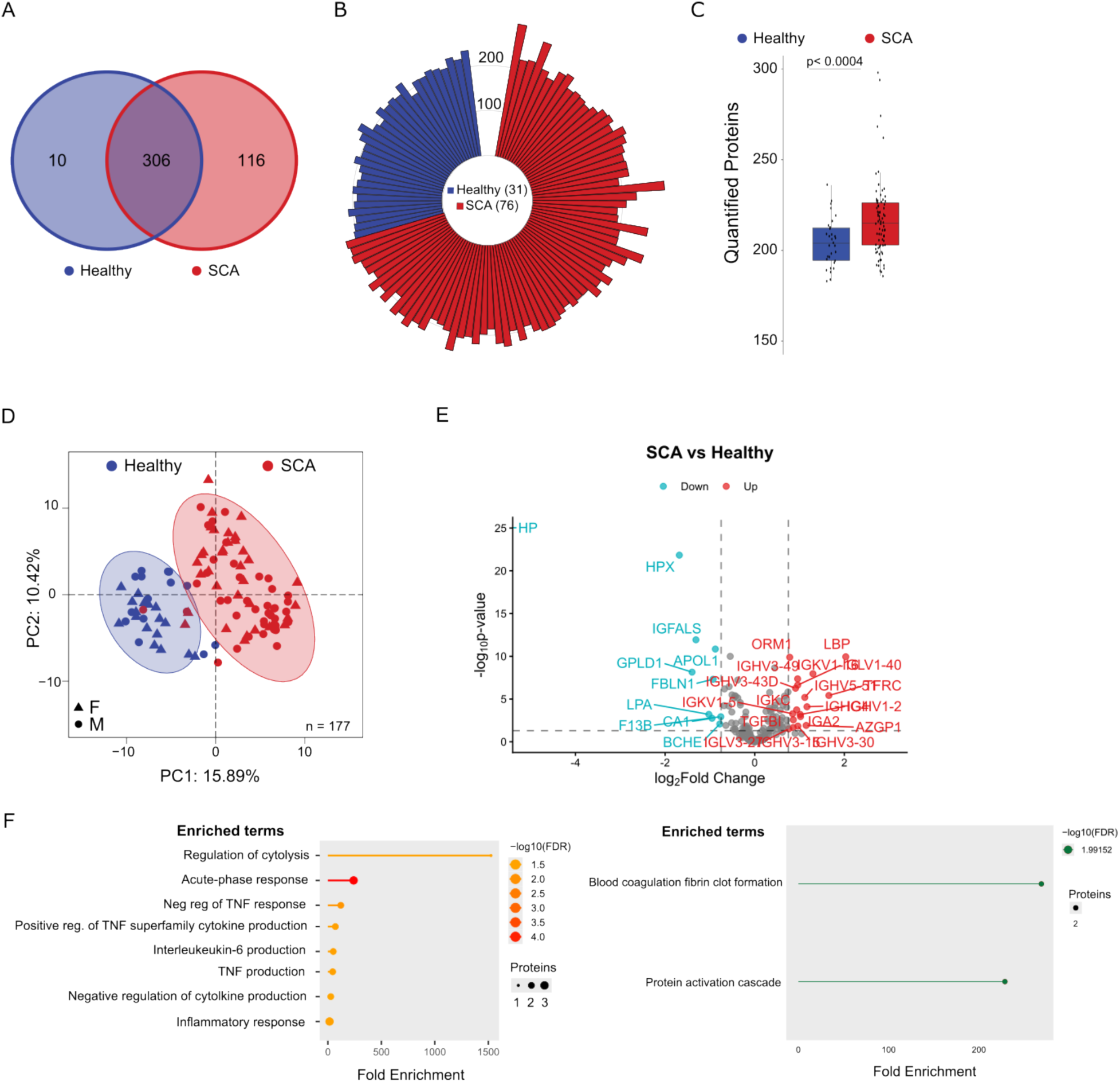
Comparative plasma proteome profiling of healthy controls (Healthy) and Sickel Cell Anaemia (SCA) patients. **A.** Overlap of total proteins quantified in healthy controls and SCA patients. **B.** Circular bar plot representing number of quantified proteins in individual samples. **C.** Box plot comparison of number of quantified proteins in healthy controls and SCA patients. **D.** Principal Component (PC) analysis using proteins quantified in all samples, showing distinct clusters for SCA patients and healthy controls. **E.** Volcano plots showing differentially abundant proteins identified in the ‘SCA vs Healthy’ comparison. Vertical dashed lines represent absolute Fold Change threshold of 0.75, while horizontal dashed lines represent p-value threshold of 0.05. **F.** Gene Ontology enrichment analysis depicting biological pathways enriched in upregulated (left) and downregulated (right) proteins using False Detection Rate (FDR) threshold of 0.05.

#### Effect of HU-treatment on the plasma proteome

Next, we analyzed the impact of HU-treatment on the plasma proteome of SCA patients. The dataset comprised 31 healthy individuals, 41 patients without (w/o)-HU treatment and 35 patients with (w/)-HU treatment (**Fig. 1**). In terms of number of quantified proteins, w/-HU patients exhibited a significantly higher heterogeneity when compared to both healthy controls and w/o-HU patients (Suppl **Fig. 3A**). Herein, we performed three comparative analyses of plasma-proteomes: 1) comparison of SCA patients without HU treatment with healthy controls (w/o-HU *vs* Healthy); 2) comparison of SCA patients on HU treatment with healthy controls (w/-HU *vs* Healthy); and, 3) comparison of SCA patients on HU treatment with those without HU treatment (w/-HU *vs* w/o-HU). PCA showed distinct clusters for both w/o- HU and w/-HU compared to healthy but had a significant overlap between them (**Fig. 3A**). The ‘w/o-HU *vs* Healthy’ analysis revealed 28 DAPs (**Fig. 3B & 3D, Supplementary Table S2**) whereas the ‘w/-HU *vs* Healthy’ comparison showed 32 DAPs (**Fig. 3C & 3D, Supplementary Table S3**). Although majority of DAPs were common in the above two analyses, there were several which were found to be differentially abundant only in either of the two analyses (**Fig. 3D**). For instance, the ‘w/o-HU vs Healthy’ comparison identified extracellular matrix protein 1 (ECM1), platelet basic protein (PPBP) and plasma protease C1 inhibitor (SERPING1) to be uniquely downregulated whereas Ficolin-3 (FCN3) among others showed unique upregulation (**Fig. 3D, Supplementary Figure 4A**). On the other hand, in the ‘w/-HU vs Healthy’ comparison, apart from significant downregulation of apolipoprotein L1 (APOL1), carbonic anhydrase 1 (CA1), glycosylphosphatidylinositol-specific phospholipase D1 (GPLD1), haptoglobin (HP), and hemopexin (HPX), we observed specific upregulation of Zinc-α-2-Glycoprotein (AZGP1), cluster of differentiation 14 (CD14), Complement component 4B (C4B) and galectin 3 binding protein (LGALS3BP) among others whereas butyrylcholinesterase (BCHE) was specifically downregulated (**Fig. 3C&D, Supplementary Figure 4B**).

**Figure 3:**
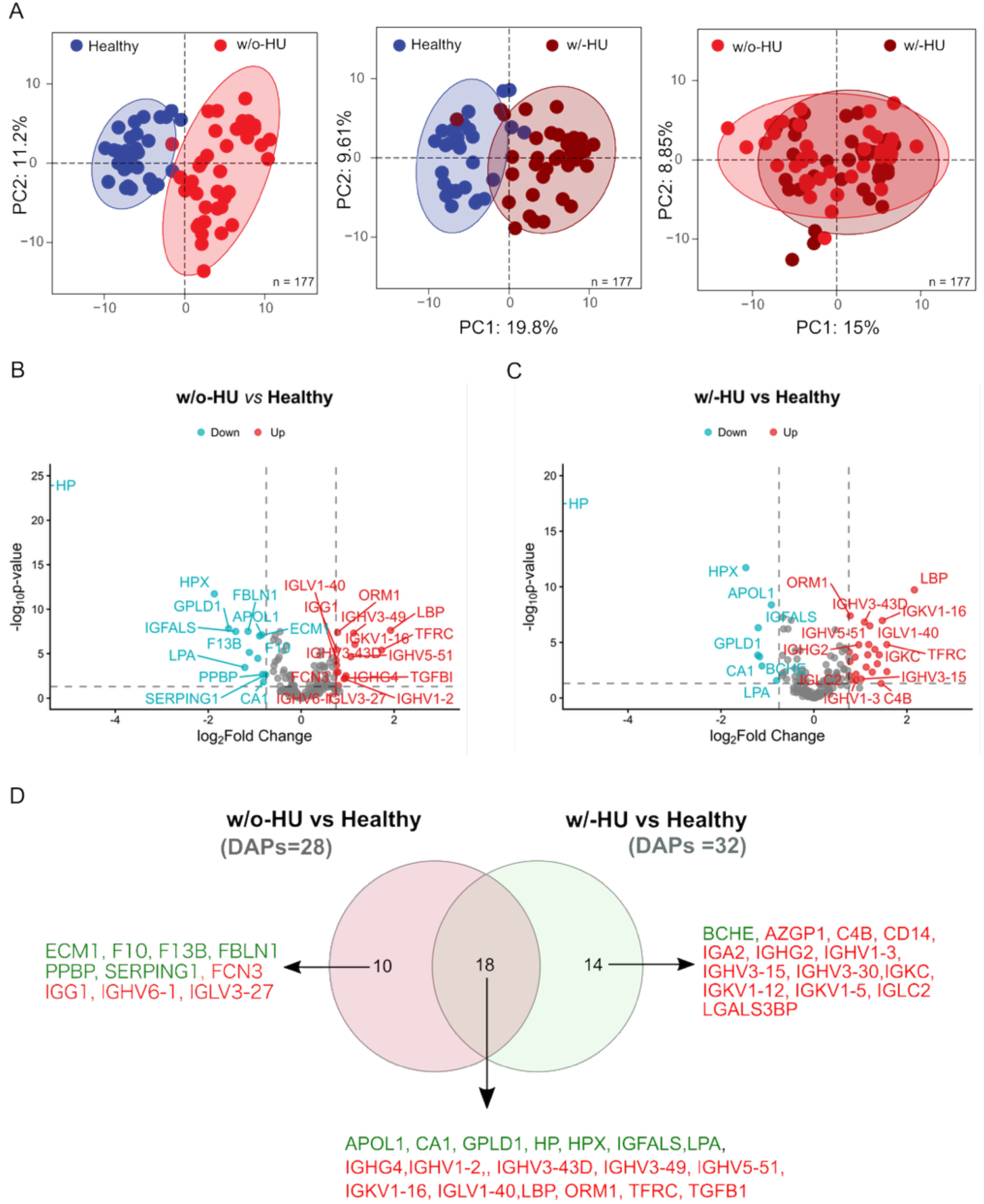
Comparative analysis of plasma proteome between healthy controls (Healthy) and Sickle Cell Anaemia (SCA) patients based on hydroxyurea (HU) treatment status. **A.** Principal Component (PC) analysis plots depicting sample clustering based on their disease and HU treatment status. **B.** Volcano plot illustrating differentially abundant proteins (DAPs) identified between patients without HU treatment (w/o-HU) and healthy controls. Vertical dashed lines represent absolute Fold Change threshold of 0.75, while horizontal dashed lines represent p-value threshold of 0.05. **C.** Volcano plot showing DAPs identified between patients with HU treatment (w/-HU) and healthy controls. Vertical dashed lines represent absolute Fold Change threshold of 0.75, while horizontal dashed lines represent p-value threshold of 0.05. **D.** Overlap of DAPs identified in ‘w/o-HU *vs* Healthy’ and ‘w/-HU *vs* Healthy’ analysis. Text in green represents downregulated proteins whereas text in red represents upregulated proteins.

The ‘w/-HU *vs* w/o-HU’ analysis showed reduced abundance of plasma immunoglobulin D (IgD) levels but increased abundance of F13B in HU-treated patients compared to those who did not receive the therapy (**Supplementary Figure 3D**). We then compared the DAPs that were common between the ‘w/o-HU vs Healthy’ and ‘w/-HU vs Healthy’ analyses. Amongst these DAPs, majority of proteins displayed largely similar abundance in w/-HU and w/o-HU samples (**Figure 4A**, **4B & 4C**). However, interestingly, HU-treatment led to relatively higher abundance of GPLD1, HP and HPX **(Figure 4B & 4C)**. *Influence of age and gender on differentially abundant proteins*

**Figure 4:**
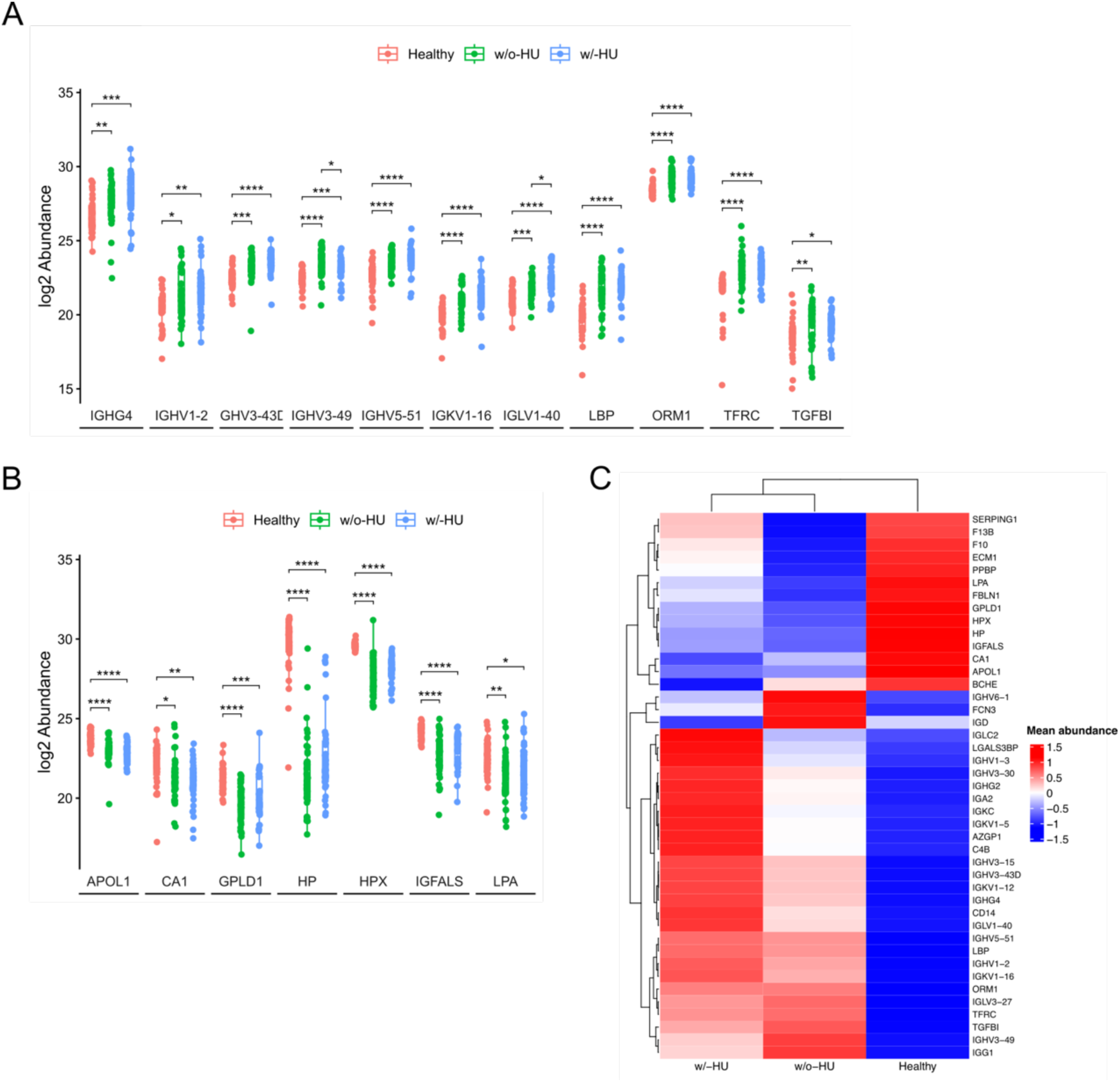
The effect of hydroxyurea (HU) treatment on the abundance of commonly deregulated plasma proteins. **A.** Box plot showing abundance of plasma proteins identified as upregulated in both ‘w/o-HU *vs* Healthy’ and ‘w/-HU *vs* Healthy’ analyses. **B.** Box plot showing abundance of plasma proteins identified as downregulated in both ‘w/o-HU *vs* Healthy’, and ‘w/-HU vs Healthy’ analyses. **C.** Heatmap showing abundance of the 43 plasma proteins identified as differentially abundant across the four sets of analyses, namely, ‘SCA *vs* Healthy’, ‘w/o-HU *vs* Healthy’, ‘w/-HU *vs* Healthy’ and ‘w/-HU *vs* w/o-HU’. *, p < 0.05; **, p < 0.01; ***, p < 0.001; ****, p < 0.0001

Overall analysis yielded 43 unique DAPs across multiple comparisons. Of these, plasma levels of 14 proteins including APOL1, AZGP1, BCHE and TFRC among others have been reported to be influenced by both age and sex in general (Niu *et al*., 2025)(**Supplementary Figure** 6A). On the other hand, 8 and 7 DAPs respectively, including C4B, CA1, ECM1, GPLD1 and TGFB1 are reported to be influenced by either age or sex (Niu *et al*., 2025)(**Supplementary Figure** 7A).

Therefore, we also assessed the abundance of DAPs in relation to gender and age. Although, overall, no discernible sex-specific differences in the plasma proteome were observed in either healthy individuals or SCA patients (**Fig. 2D**), a few DAPs showed sex-specific effects. Amongst the upregulated proteins, for Immunoglobulin A, Alpha Heavy Chain 2 (IGA2) and IGHV3-30 and Immunoglobulin lambda variable 3-27 (IGLV3-27), significant upregulation was observed only in female patients whereas for Immunoglobulin kappa constant (IGKC), Immunoglobulin heavy constant gamma 4 (IGHG4), Immunoglobulin kappa variable 1-5 (IGKV1-5) and TGFBI, were upregulated specifically in male patients (**Fig. 5A**). Similar observations were made for FCN3, IGHV1-2, IGLV3-27 and TGFBI in the ‘w/o-HU vs Healthy’ comparison (**Supplementary Figure 5A**). In addition, we also observed gender specific upregulation of CD14, IGHG2, IGHG4, IGKC, and LGALS3BP in ‘w/-HU vs Healthy’ analysis (**Supplementary Figure 5B**).

**Figure 5:**
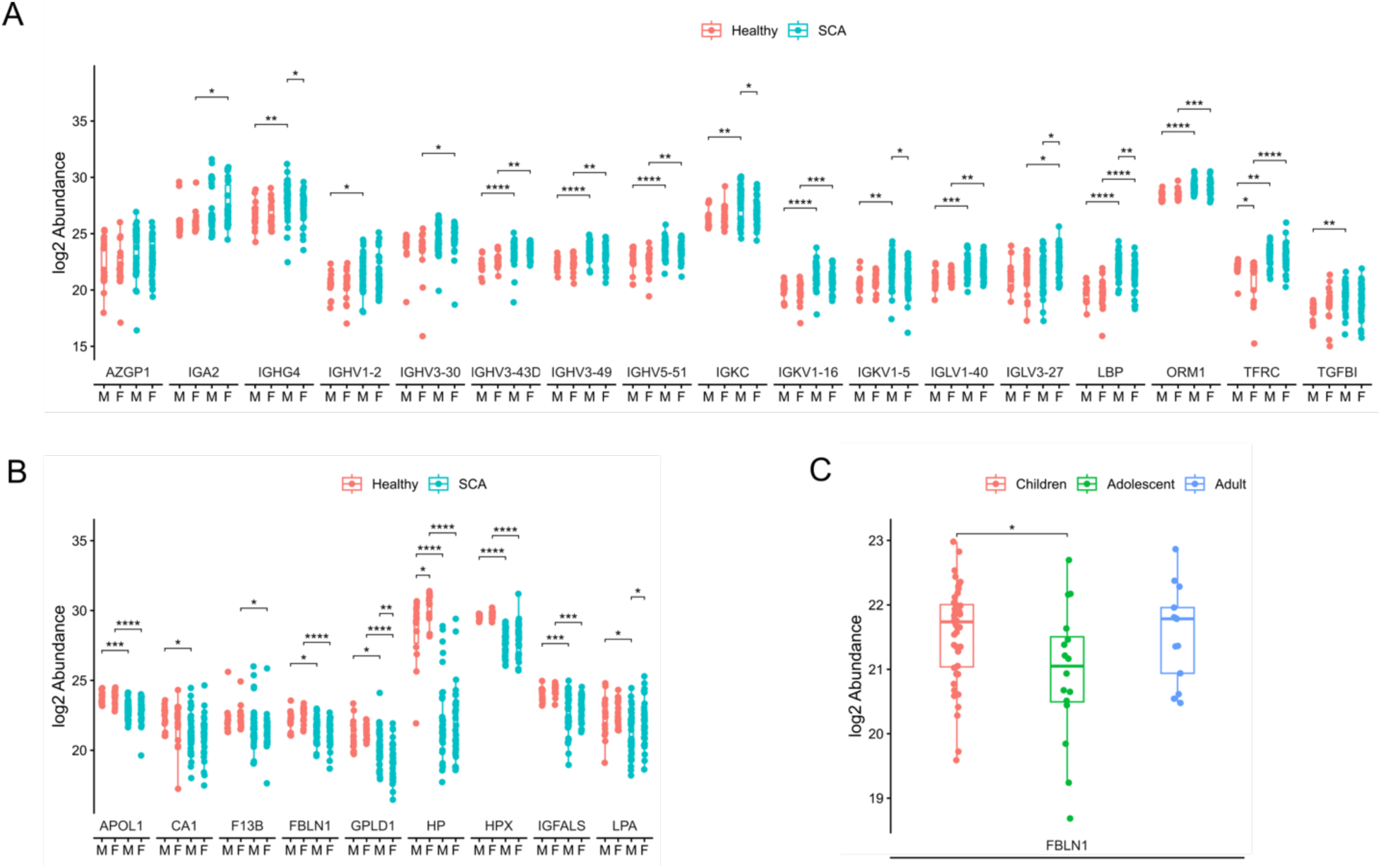
Quantitative representation of Differentially Abundant Proteins (DAPs) identified in ‘SCA *vs* Healthy’ analysis in SCA patients and healthy controls across gender and age groups. **A.** Box plot showing gender-wide abundance of upregulated proteins. **B.** Box plot showing gender-wise abundance of downregulated proteins. **C.** Box plot showing age-group-wise abundance of downregulated proteins in SCA patients. *, p < 0.05; **, p < 0.01; ***, p < 0.001; ****, p < 0.0001 M – Male; F – Female; SCA – Sickle Cell Anaemia

For the downregulated proteins, F13B downregulation was observed only in females whereas CA1 and Lipoprotein A (LPA), showed downregulation only in males (**Fig. 5B**). Additionally, glycosylphosphatidylinositol-specific phospholipase D (GPLD1) downregulation was relatively higher in female patients compared to males. Haptoglobin (HP) levels were significantly elevated in healthy females than males but similar in male and female patients, indicating a stronger downregulation in female patients (**Fig. 3A**). The ‘w/o-HU vs Healthy’ comparison revealed specific downregulation of PPBP in male patients (**Supplementary Figure 5A**). Similarly, the ‘w/-HU vs Healthy’ analysis displayed specific downregulation of BCHE, CA1, and LPA in male patients (**Supplementary Figure 5B**).

Age-group analysis revealed that while none of the upregulated proteins displayed any age-specific bias, downregulation of FBLN1 was more in adolescent patients than in children or adults (**Fig. 3C**). Similar observations were made for IGHV6-1 and FBLN1 in the ‘w/o-HU vs Healthy’ analysis (**Supplementary Fig. 3B**). Differential abundance of BCHE, CD14, GPLD1 and LGALS3BP in patients of different age groups was also observed in the ‘w/-HU vs Healthy’ analysis (**Supplementary Fig. 3C**).

#### Overlap with reported signatures in beta thalassemia

With the aim to identify distinct repertoires of DAPs present in the plasma of SCA patients, we compared them with those reported to be differentially abundant between beta-thalassemia patients. Out of the 43 unique DAPs identified across subgroup analyses in this study, 23 including APOL1, AZGP1, CA1, FBLN1, HP, HPX, ORM1, and TFRC, exhibited overlap with those differentially abundant in beta-thalassemia patients (Li *et al*., 2022a). Further, 10 proteins, such as F10, LPA, FCN3, and others, were common to those showing differential abundance in beta-thalassemia patients after HU treatment. However, twenty-two proteins, including C4B, LGALS3BP, BCHE, SERPING1, F13B, LBP, and ECM1, demonstrated unique differential abundance in SCA (**Supplementary Figure 7B**).

#### Global correlation and protein-protein network analysis

A comprehensive global correlation analysis of the 43 DAPs identified two distinct clusters of highly correlated proteins (**Figure. 6A**). Subsequently, STRING analysis of the 43 DAPs revealed a robust protein interaction network, wherein majority of the proteins were interconnected (**Figure. 6B**).

**Figure 6:**
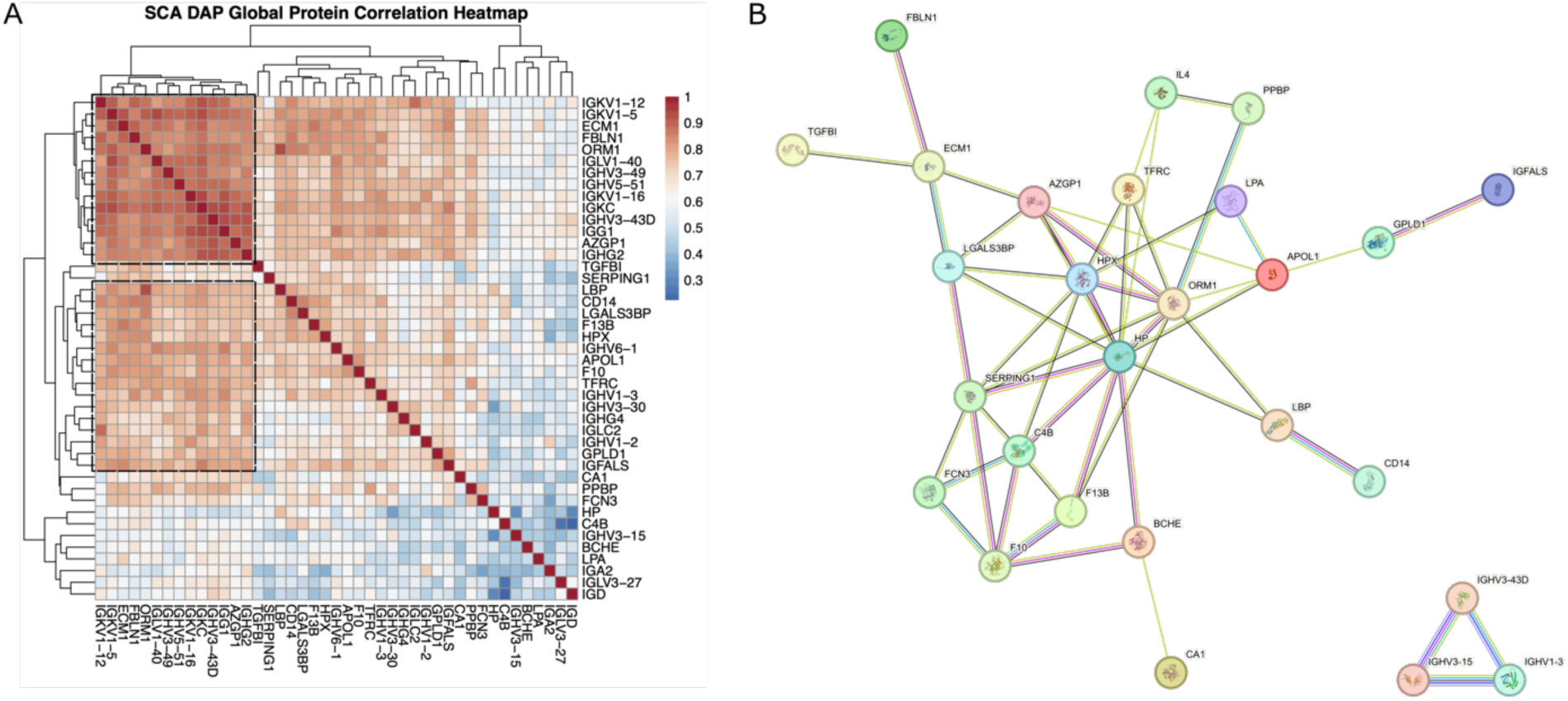
Global correlation and protein-protein interaction network analysis of the 43 Differentially Abundant Proteins (DAPs) identified in the study. **A.** Heatmap representing correlation amongst the 43 DAPs. Dashed line boxes indicate identified clusters. **B.** Search Tool for the Retrieval of Interacting Genes/Proteins (STRING) network analysis for the 43 DAPs.

#### Correlation analysis of protein abundance to clinical phenotype/parameters

To further evaluate the clinical relevance of the differentially abundant proteins, we correlated their abundance with various clinical phenotypes and parameters (**Table 3**). At a nominal significance level of p <0.05, abundance of several critical proteins, including APOL1, AZGP1, FBLN1, GPLD1, HPX, LGALS3BP, TFRC correlated with multiple parameters, such as frequency of blood transfusions and VOC, WBC and platelet count, and others like levels of serum albumin and ALT which are indicators of liver function. Consistent with the nature of the disease, HPX abundance exhibited a moderate but significant positive correlation with RBC counts while demonstrating a negative correlation with HbS levels, reticulocyte counts and serum bilirubin levels. Additionally, GPLD1 abundance correlated with serum albumin, urine-microalbumin and ALT levels. Furthermore, TGFBI abundance displayed correlation with neutrophil and leukocyte counts.

**Table 3:** Correlation of differentially abundant proteins with clinical phenotypes and parameters.

| Clinical Phenotype/ Parameter | Differentially Abundant Proteins |
| --- | --- |
| AFI-Infection | GPLD1, IGA2, IGD |
| Blood transfusion (BT) | APOL1, FBLN1, GPLD1, IGFALS, AZGP1, IGA2, IGHG4, IGHV1_2, IGHV3_30, IGHV3_43D, IGHV3_49, IGHV5_51, IGKC, IGKV1_16, IGKV1_5, IGLV3_27, LBP, ORM1, TFRC, ECM1, PPBP, SERPING1, FCN3, IGG1, IGHG2, IGLC2, LGALS3BP, IGD |
| Vaso-occlusive crisis (VOC) | APOL1, F13B, GPLD1, IGFALS, AZGP1, IGA2, IGHG4, IGHV1_2, IGHV3_43D, IGKC, IGKV1_5, IGLV3_27, LBP, ORM1, TFRC, FCN3, CD14, IGHG2, IGLC2 |
| RBC | HPX, IGHV1-3 |
| WBC | F13B, FBLN1, GPLD1, HPX, IGFALS, AZGP1, IGHV3-15, IGHV3-30, IGHV3-43D, IGHV5-51, IGKC, IGKV1-5, IGLV1-40, IGLV3-27, ECM1, PPBP, SERPING1, FCN3, IGG1, CD14, IGHG2, IGHV1-3, IGLC2, LGALS3BP |
| Platelets | APOL1, F13B, FBLN1, GPLD1, HP, HPX, IGFALS, AZGP1, IGHG4, IGHV1-2, IGHV3-15, IGHV3-30, IGHV3-43D, IGHV3-49, IGHV5-51, IGKC, IGKV1-16, IGKV1-5, IGLV1-40, IGLV3-27, LBP, ORM1, ECM1, F10, PPBP, SERPING1, FCN3, IGG1, IGHV6-1, C4B, CD14, IGHG2, IGHV1-3, IGKV1-12, IGLC2, LGALS3BP |
| Neutrophils | TGFBI, PPBP |
| Leukocytes | TGFBI |
| Eosinophils | AZGP1, IGHV3-43D |
| Reticulocyte | FBLN1, HPX, IGFALS, CD14 |
| Mean corpuscular hemoglobin (MCH) | APOL1, IGHV1-2, IGLV3-27, PPBP, FCN3 |
| Mean corpuscular hemoglobin concentration (MCHC) | FCN3 |
| Mean corpuscular volume (MCV) | IGLV3-27, PPBP |
| HbA | APOL1, BCHE, CA1, FBLN1, HP, IGFALS, AZGP1, IGHG4, IGKC, IGKV1-16, IGKV1-5, IGLV1-40, LBP, ORM1, TFRC, TGFBI, ECM1, F10, IGG1, IGHV6-1, C4B, CD14, IGHG2, IGLC2, LGALS3BP |
| HbS | APOL1, BCHE, F13B, HP, HPX, IGFALS, LBP, ORM1, ECM1, FCN3, CD14, IGHV1-3, LGALS3BP |
| Serum-albumin | APOL1, BCHE, CA1, F13B, FBLN1, GPLD1, HP, HPX, IGFALS, LPA, AZGP1, IGA2, IGHG4, IGHV1-2, IGHV3-15, IGHV3-30, IGHV3-43D, IGHV3-49, IGHV5-51, IGKC, IGKV1-16, IGKV1-5, IGLV1-40, IGLV3-27, LBP, ORM1, TFRC, TGFBI, ECM1, F10, PPBP, SERPING1, FCN3, IGG1, IGHV6-1, C4B, CD14, IGHG2, IGHV1-3, IGKV1-12, IGLC2, LGALS3BP, IGD |
| Serum-Bilirubin | FBLN1, HPX, IGHV1-2, IGHV3-15 |
| Urine-microalbumin | CA1, FBLN1, GPLD1, IGFALS, AZGP1, IGA2, IGHG4, IGHV3-30, IGHV3-43D, IGHV3-49, IGHV5-51, IGKC, IGKV1-5, IGLV1-40, ORM1, TFRC, F10, SERPING1, IGG1, CD14, IGHG2, IGHV1-3, IGKV1-12, IGLC2, LGALS3BP, IGD |
| Alanine aminotransferase (ALT) | APOL1, CA1, F13B, FBLN1, GPLD1, HPX, IGFALS, LPA, AZGP1, IGA2, IGHG4, IGHV1-2, IGHV3-15, IGHV3-30, IGHV3-49, IGHV5-51, IGKC, IGKV1-16, IGKV1-5, IGLV3-27, ORM1, TFRC, ECM1, F10, PPBP, IGG1, IGHV6-1, CD14, IGHG2, IGKV1-12, IGLC2, IGD |
| Alkaline Phosphatase (ALP) | APOL1, GPLD1, IGFALS, AZGP1, IGA2, IGHG4, IGHV1-2, IGHV3-30, IGHV3-43D, IGHV3-49, IGHV5-51, IGKC, IGKV1-16, IGKV1-5, IGLV1-40, LBP, ORM1, TFRC, TGFBI, ECM1, PPBP, IGG1, IGHV6-1, CD14, IGHG2, IGKV1-12, IGLC2, LGALS3BP, IGD |

## Discussion

The clinical manifestations of the single-gene disease SCA are highly heterogeneous and depend on interactions with other genetic and environmental factors (Habara and Steinberg, 2016; Jain and Mohanty, 2018; Inusa *et al*., 2019). SCA manifests at a very young age, typically within a few years of birth in majority of patients but the life expectancy has risen to ∼ 50 years with the aid of modern healthcare (Tanabe *et al*., 2019). A recent study reported longer lifespan in female SCA-patients suggesting a gender-specific disease manifestation and life expectancy (Jiao *et al*., 2023). Severity of clinical symptoms also vary and partly depend on the response to HU-therapy (Charache *et al*., 1995; Thornburg *et al*., 2012; McGann and Ware, 2015; Pandey *et al*., 2022; George *et al*., 2025). The mechanistic understanding of the clinical heterogeneity in SCA is currently limited and identification of robust biomarkers with predictive potential is an area of intense research.

In this study, we present a comparative plasma proteome map of Indian SCA patients and healthy individuals. Our analysis revealed significant differences in protein abundances, with SCA patients exhibiting a more heterogeneous plasma proteome. Proteins involved in several key pathway including blood coagulation, proteolysis, immune regulation, and inflammation, related to SCA pathophysiology were found to be deregulated. Functionally, the downregulation of proteins involved in coagulation and proteolysis may contribute to increased risk of thrombotic events and exacerbate the pathological consequences of the disease. Conversely, the upregulation of immune response-related proteins associated with inflammation and immune activation is likely attributed to chronic hemolysis and vaso-occlusion, two critical features of SCA. For example, Transforming growth factor beta induced (TGFBI) is upregulated in SCA patients. Interestingly, transforming growth factor beta (TGF-β) plays a key role in SCD pathogenesis, affecting endothelial and vascular dysfunction, inflammation, and hematopoietic homeostasis (Nolan *et al*., 2006; Frangogiannis, 2017; Santiago *et al*., 2020, 2021). In this regard, we also observed correlation of TGFBI abundance with neutrophil and leukocyte counts. Both, adult and pediatric SCD patients are reported to have plasma levels of TGF-β1 higher than the control group (Nolan *et al*., 2006; Frangogiannis, 2017; Santiago *et al*., 2020, 2021). Moreover, in the latter study, in SCA patients, TGF-β1 levels positively correlated with red blood cells, hemoglobin, hematocrit, platelets, and TIMP-1, suggesting its role in vascular remodeling, vasculopathy, angiogenesis, and inflammation. APOL1, which we observed to be downregulated in SCA-patients, is also reported to be downregulated in beta-thalassemia patients (Li *et al*., 2022b). Interestingly, genetic variants in *APOL1* are implicated in the development and progression of SCD-related nephropathy and albuminuria (Freedman and Skorecki, 2014; Zahr *et al*., 2019; Ataga, Saraf and Derebail, 2022). Importantly, we also observed correlation of APOL1 abundance with multiple parameters including serum albumin levels. Fibulin-1 (FBLN1) is an extracellular glycoprotein that can modulate cell morphology, growth, adhesion and motility and dysregulation of certain FBLNs occurs in a range of human cancers, such as prostate cancer (Gallagher, Currid and Whelan, 2005). It is found in many extracellular matrices including those of blood vessels. It is abundant in blood and its reduced levels in circulation are correlated with early-onset atherosclerosis (Nissinen *et al*., 2009), which in turn might be a risk factor for stroke in SCA.

A substantial body of evidence supports the influence of sex and age on the plasma proteome (Koprulu *et al*., 2025; Niu *et al*., 2025). Furthermore, numerous studies have documented sex and age-related variations in complications associated with SCA. Several proteins, including GPLD1, HP and LPA were differentially regulated in male and female SCA patients in this study. GPLD1 regulates glucose and lipid metabolism, and is implicated in several chronic diseases (Cao *et al*., 2023). Importantly, downregulation of genes including GPLD1 and APOC1 that are involved in triglyceride and neutral lipid metabolism has also been reported in beta-thalassemia patients (Li *et al*., 2022b). HP is the plasma protein with highest binding affinity for Hb. Upon RBC lysis, the free hemoglobin is sequestered by haptoglobin. This HP-Hb complex is subsequently cleared by macrophages in liver and spleen. However, in severe hemolysis, haptoglobin levels may become depleted. We observed relatively increased abundance of HP in hydroxyurea treated patients likely indicating the role of hydroxyurea in preventing hemolysis. Further, it has been shown that in Hp^-/-^ mice, there is increased deposition of Hb in proximal tubules of kidney (Shih, McFarlane and Verhovsek, 2014; Abah *et al*., 2018). Similarly, LPA is a complex molecule consisting of a low-density lipoprotein (LDL) particle that is covalently linked to apolipoprotein (a). In SCA, due to hemolysis, there is rapid turnover of RBC and thus increased demand for reticulocyte membranes. On the other hand, upregulated proteins like AZGP1 are known in lipid metabolism and inflammation.

A comprehensive analysis of patients with and without HU treatment yielded intriguing results. In the comparison between without-HU and Healthy groups, proteins such as ECM1, F10, and F13B, PPBP, SERPING1, and TTR exhibited unique downregulation. Conversely, FCN3 and IGG1, along with other immunoglobulins, were upregulated. ECM1 is a secretory glycoprotein shown to promote angiogenesis by stimulating the proliferation of endothelial cells (Han *et al*., 2001). Notably, F10 (Factor X) is a crucial enzyme in the blood coagulation cascade. Additionally, PPBP (Pro-Platelet Basic Protein), also known as CXCL7 (Chemokine Ligand 7) and F13B (Factor XIII Subunit B) also play a key role in blood clot formation and stabilization respectively.

In addition, Hydroxyurea treatment, a common therapy for SCA, significantly impacted the plasma, with varying effects on protein abundance depending on treatment status and patient characteristics. In the comparison between patients with-HU and Healthy groups, BCHE was uniquely downregulated whereas AZGP1, C4B and CD14 among others showed unique upregulation. BCHE is primarily synthesized in liver and is known to decrease in acute and chronic liver injury, cirrhosis, malnutrition, and in response to stress and inflammation (Lampón *et al*., 2012; Ramachandran *et al*., 2014). It is also reported to be downregulated in beta-thalassemia patients who respond to HU treatment compared to those who do not (Zohaib *et al*., 2019). AZGP1 found to be upregulated in several analyses, is a secreted glycoprotein involved in lipid metabolism, immune regulation, and inflammation both of which may play a role in the pathophysiology of sickle cell anemia (Alenad *et al*., 2022).

Overall, our results suggest a stochastic deregulation of SCA plasma proteome as indicated by the number and identity of individual proteins quantified across SCA-patients. Many of the deregulated proteins identified in our study including APOL1, AZGP1, CA1, FBLN1, FCN3, HP, HPX, ORM1, TFRC, LPB, CA1, PPBP and others have been reported to have altered levels in Beta-thalassemia patients. This observation is particularly significant because beta-thalassemia shares several pathophysiological features with SCA and the presence of similar patterns of protein deregulation in both conditions underscores the overlapping mechanisms underlying their clinical manifestations and highlights the potential for shared therapeutic targets and biomarker development. Consequently, our findings, not only corroborate the role of several proteins as biomarkers in SCA pathophysiology but also identify several potential candidate biomarkers. We do not observe large differences in plasma proteome of SCA patients with and without hydroxyurea treatment. This could likely be due to varying duration of HU therapy or differential response to HU therapy. These aspects need to be further evaluated in subsequent studies.

## Data Availability

The raw data has been uploaded in the PRIDE (https://www.ebi.ac.uk/pride) partner repository of the ProteomeXchange Consortium (https://www.proteomexchange.org), with the dataset identifier PXD062365. All the processed data are contained in the manuscript.

## Acknowledgement

The authors express their deep sense of gratitude to all the patients and healthy volunteers who consented to participate in this study.

## Ethics statement

The Institutional Ethics Committee (IEC) of CSIR-Centre for Cellular and Molecular Biology (CSIR-CCMB), Hyderabad, reviewed the proposal “CSIR-Sickle Cell Anaemia Mission” and gave ethical approval for this work. The IEC certificate number is IEC-65-R1/2019.

