## Supplementary Figures for "Plasma proteome signatures in sickle cell anaemia and effect of hydroxyurea treatment"

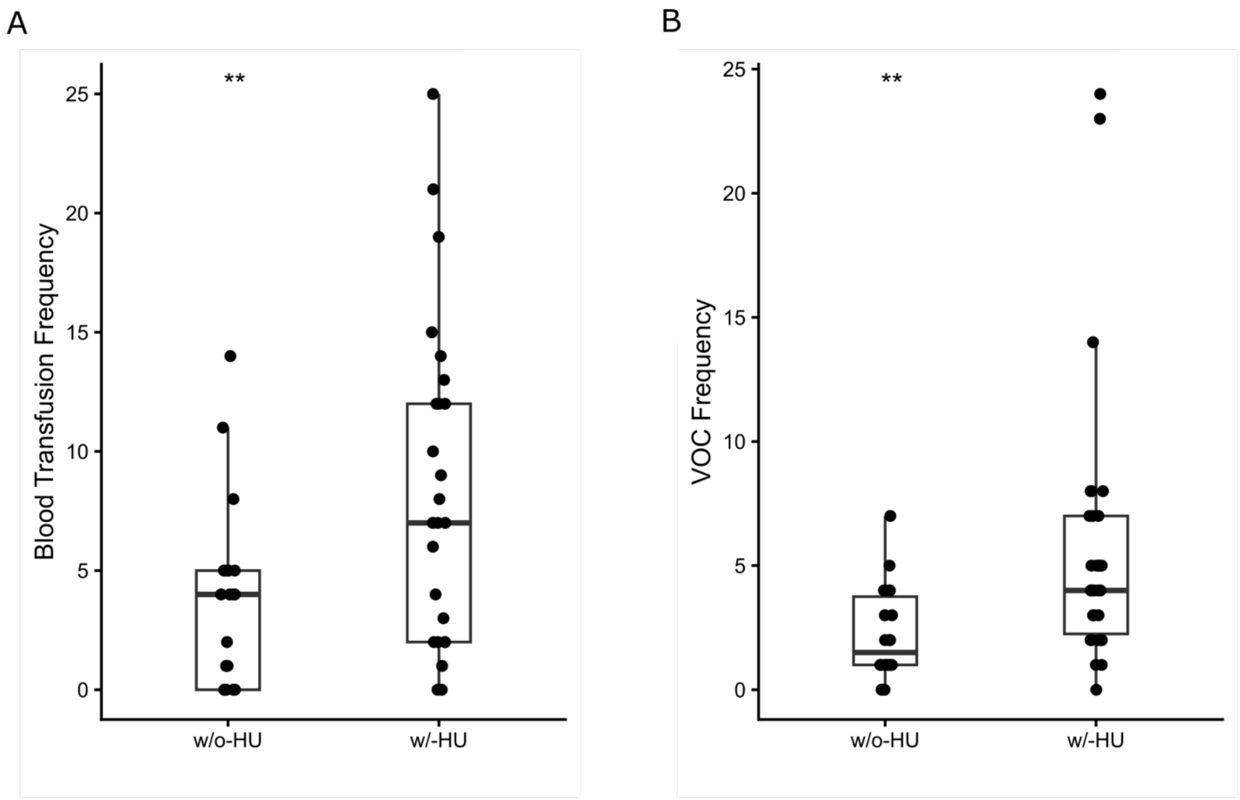


**Supplementary Figure 1:** Clinical characteristics of sickle cell anaemia (SCA) patients with hydroxyurea treatment (w/-HU) and without hydroxyurea treatment (w/o-HU).

- **A.** Box plot representing frequency of past episodes of blood transfusion in SCA patients w/o-HU and w/-HU treatment.
- **B.** Box plots representing frequency of past episodes of vaso-occlusive crisis (VOC) in SCA patients w/o-HU and w/-HU treatment.

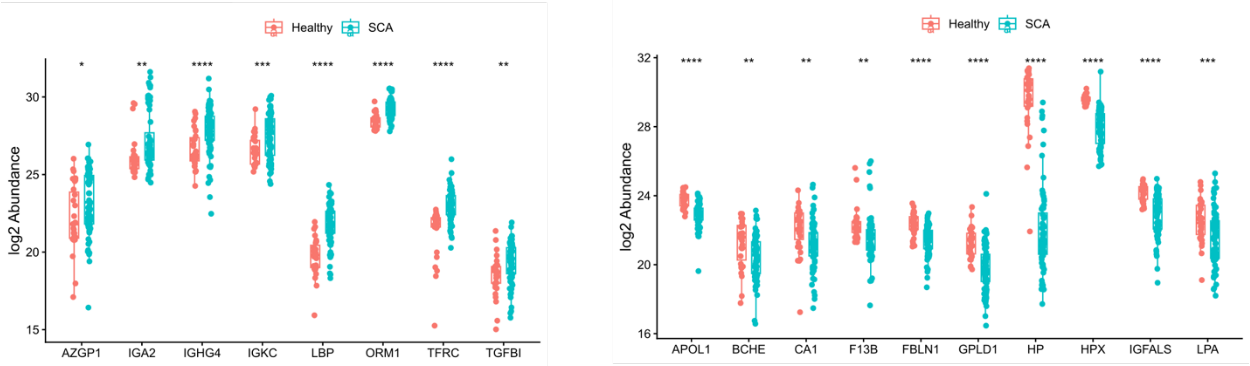


**Supplementary Figure 2:** Comparative plasma proteome analysis of sickle cell anaemia (SCA) patients and healthy controls (Healthy).

- **A.** Box plot representing abundance of proteins identified as upregulated proteins in 'SCA vs Healthy controls' analysis.
- **B.** Box plot representing abundance of proteins identified as downregulated proteins in 'SCA vs Healthy controls' analysis.

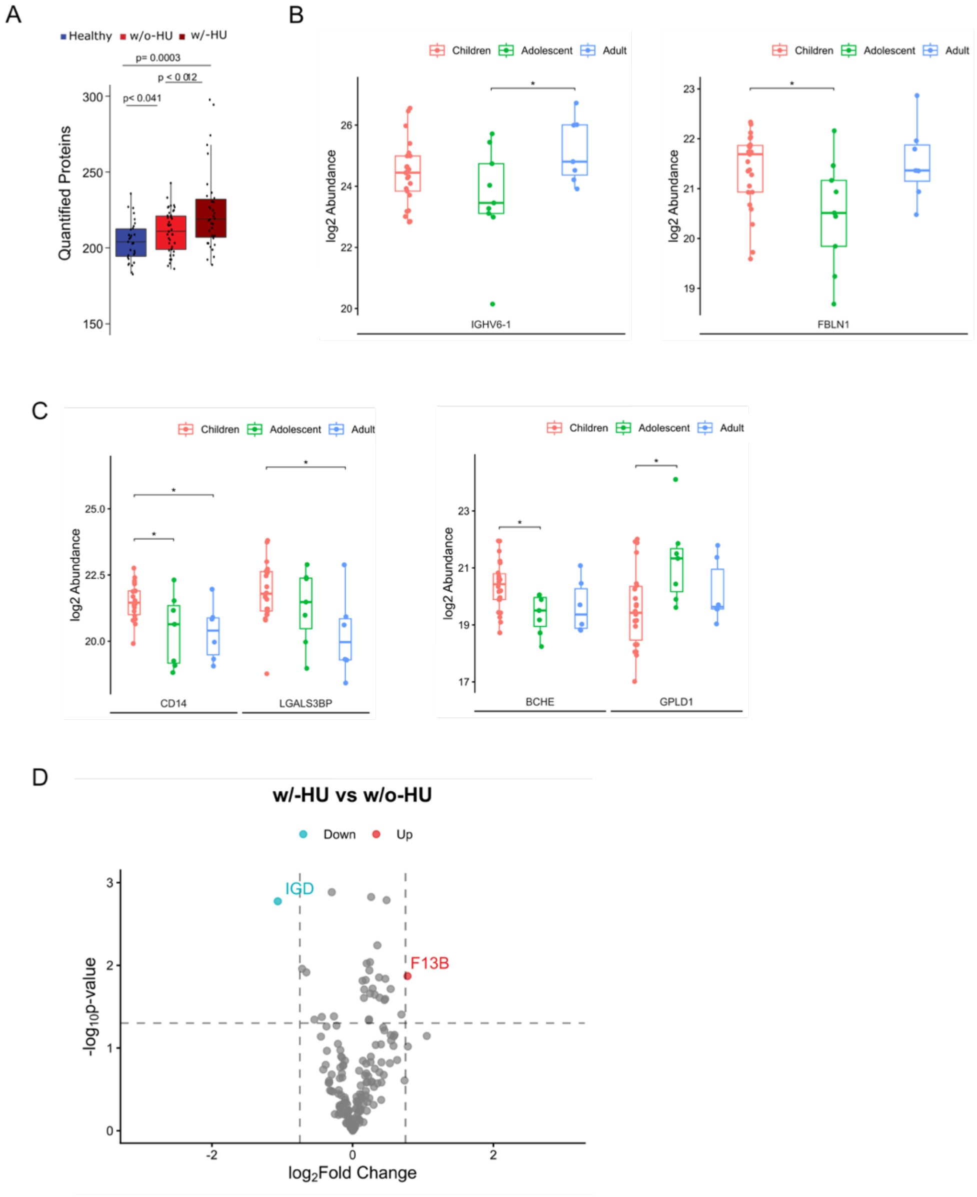


**Supplementary Figure 3:** Absolute and relative quantification of proteins in healthy controls (Healthy) and sickle cell anaemia (SCA) patients based on their hydroxyurea (HU) treatment status and age.

- **A.** Box plot showing number of quantified proteins in healthy controls (Healthy), SCA patients with HU (w/-HU) treatment, and SCA patients without HU (w/o-HU) treatment.
- **B.** Box plot for upregulated (left) and downregulated (right) proteins in 'w/o-HU vs Healthy' comparison displaying differential abundance across age groups.
- **C.** Box plot for upregulated (left) and downregulated (right) proteins in 'w/-HU vs Healthy' comparison displaying differential abundance across age groups.
- **D.** Volcano plots showing differentially abundant proteins (DAPs) in the 'w/-HU vs w/o-HU' comparison.

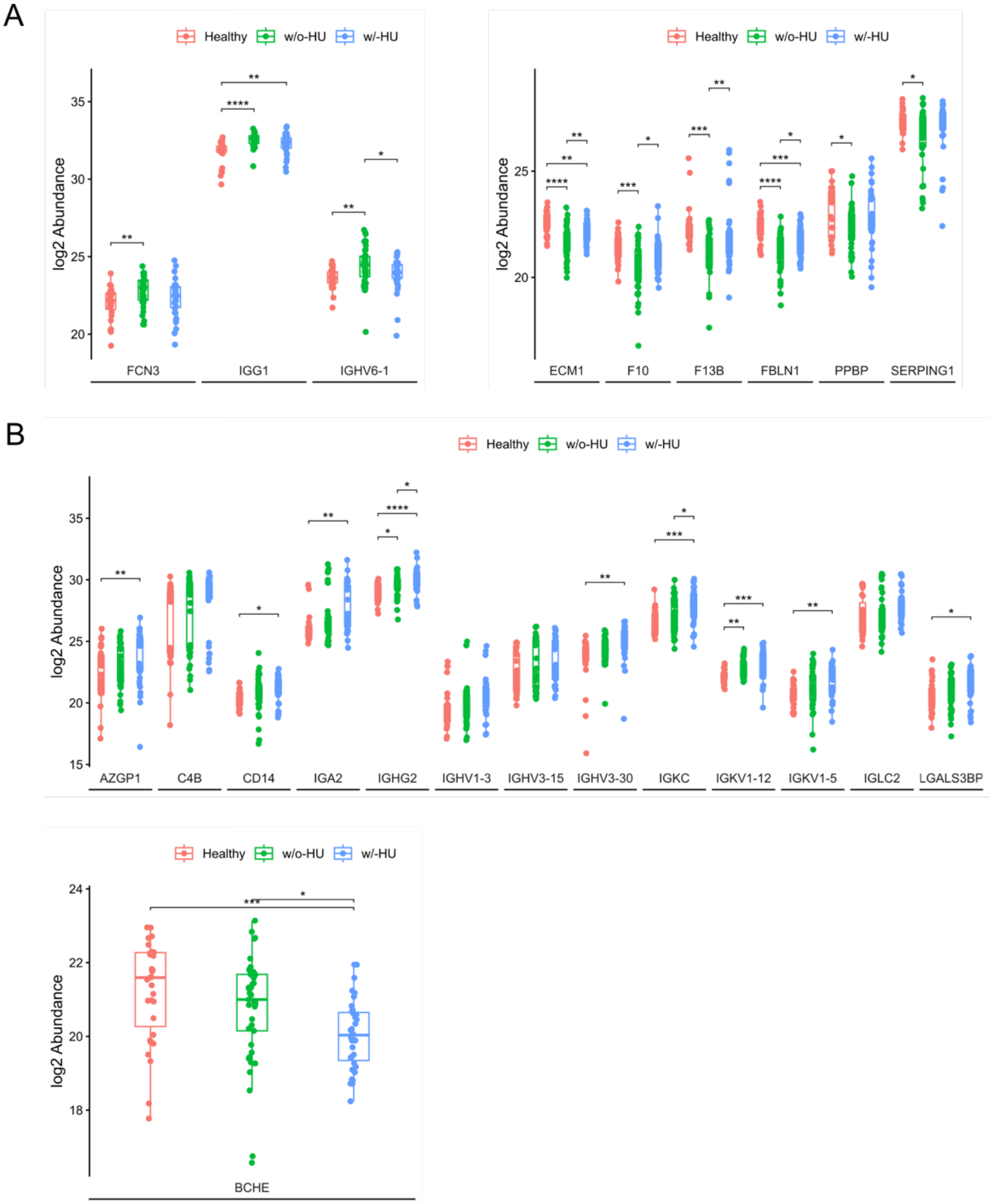


**Supplementary Figure 4:** Proteins exhibiting differential abundance only in either ‘w/o-HU vs Healthy’ or ‘w/-HU vs Healthy’ comparison.

- **A.** Box plot showing abundance of plasma proteins upregulated (left) or downregulated (right) only in ‘w/o-HU vs Healthy’ comparison.
- **B.** Box plot showing abundance of plasma proteins upregulated (top) or downregulated (bottom) only in ‘w/-HU vs Healthy’ comparison.

*, $p < 0.05$; **, $p < 0.01$; ***, $p < 0.001$; ****, $p < 0.0001$

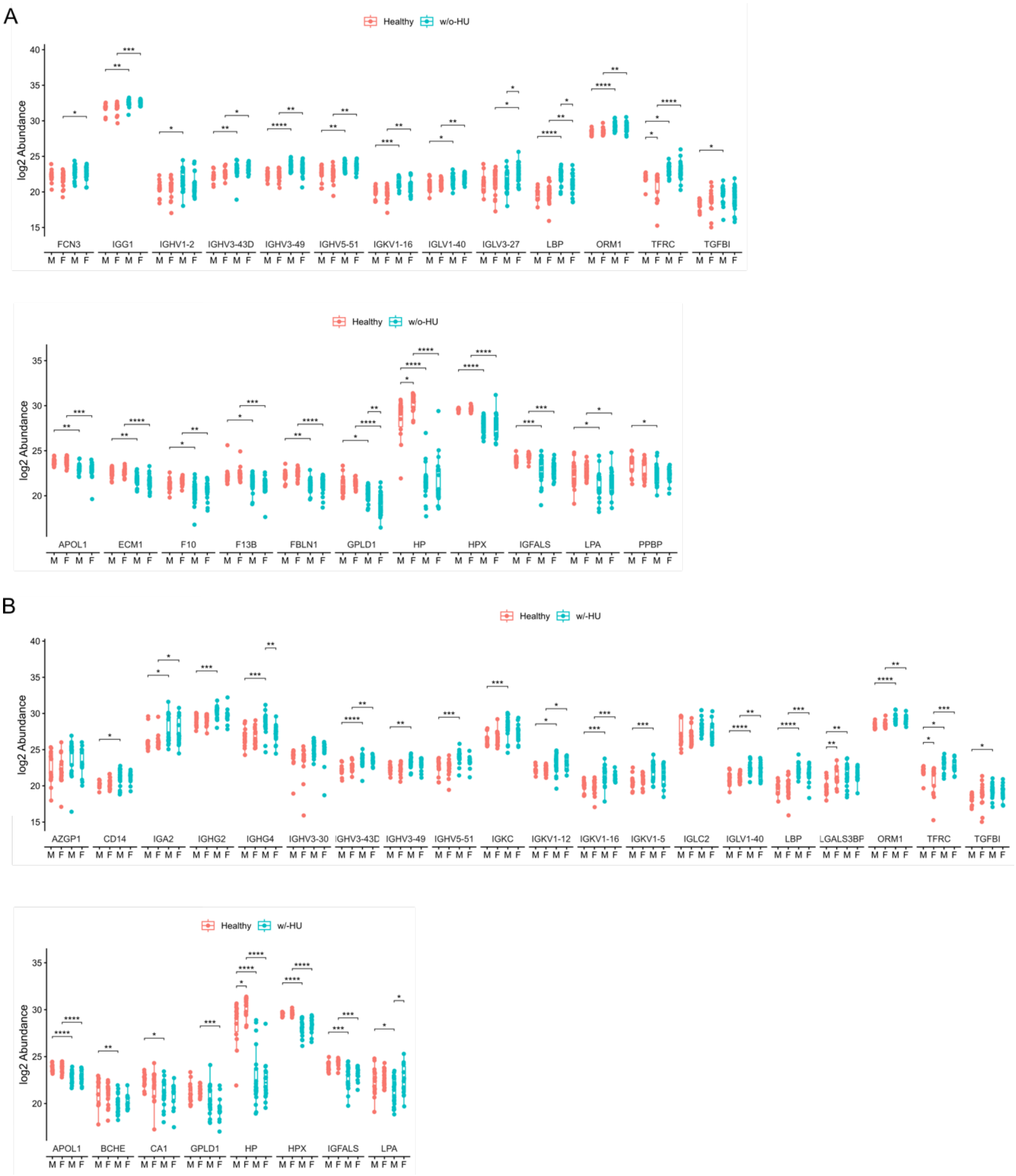


**Supplementary Figure 5: Influence of age and gender on abundance of DAPs identified in without-HU (w/o-HU) vs Healthy and with-HU (w/-HU) vs Healthy comparison.**

**A.** Box plot displaying gender-wise abundance of upregulated (top) or downregulated (down) proteins in 'w/o-HU vs Healthy' comparison.

**B.** Box plot displaying gender-wise abundance of upregulated (top) or downregulated (down) proteins in 'w/-HU vs Healthy' comparison.

*, $p < 0.05$; **, $p < 0.01$; ***, $p < 0.001$; ****, $p < 0.0001$

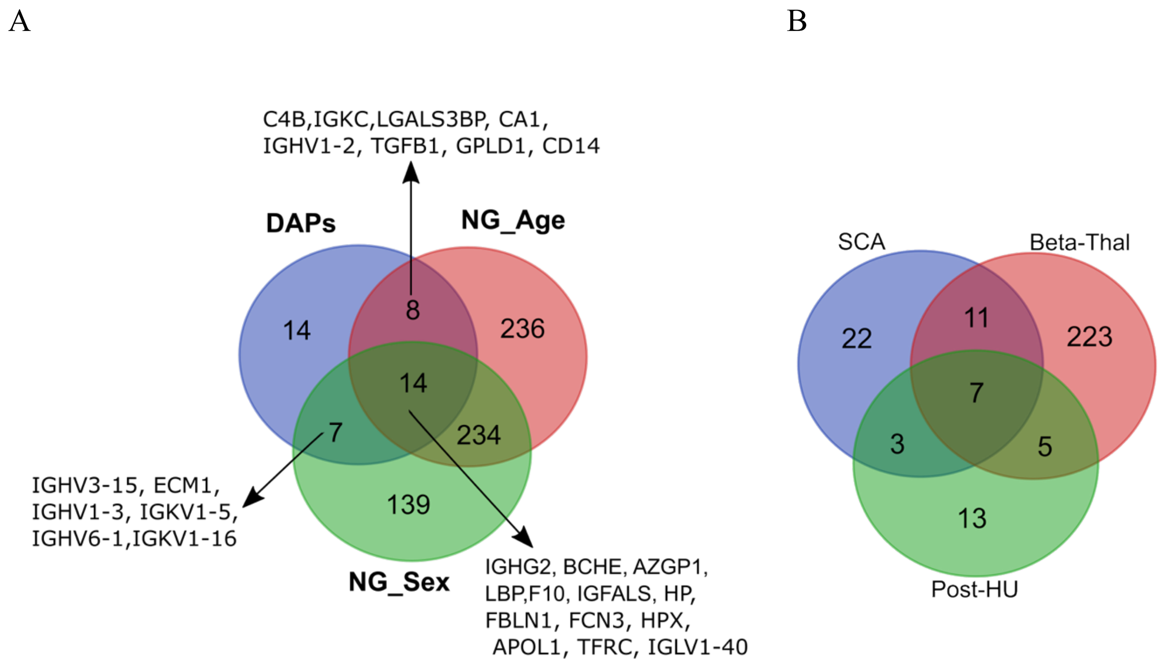


**Supplementary Figure 6:** Overlap of differentially abundant proteins (DAPs) identified in sickle cell anaemia (SCA) patients in this study with those previously published studies

A. Venn diagram illustrating overlap of DAPs with plasma proteins reported to be influenced by age and sex (Niu, L. et al. (2025))

B. Venn diagram displaying overlap of DAPs with proteins reported to be differentially abundant in beta-thalassemia (Beta-Thal) patients without hydroxyurea treatment (Li *et al.*, 2022a) or after Hydroxyurea (Post-HU) treatment (Zohaib *et al.*, 2019).
